# A distinct cognitive profile in individuals with 3q29 deletion syndrome

**DOI:** 10.1101/2021.03.05.21252967

**Authors:** Cheryl Klaiman, Stormi Pulver White, Celine Saulnier, Melissa Murphy, Lindsey Burrell, Joseph Cubells, Elaine Walker, The Emory 3q29 Project, Jennifer Gladys Mulle

## Abstract

**Background:** 3q29 deletion syndrome is associated with mild to moderate intellectual disability. However, a detailed understanding of the deletion’s impact on cognitive ability is lacking. The goal of this study was to address this knowledge gap. A second goal was to ask whether the cognitive impact of the deletion predicted psychopathology in other domains.

**Methods:** We systematically evaluated cognitive ability, adaptive behavior, and psychopathology in 32 individuals with the canonical 3q29 deletion using gold-standard instruments and a standardized phenotyping protocol.

**Results:** Mean FSIQ was 73 (range 40-99). Verbal subtest score (mean 80, range 31-106) was slightly higher and had a greater range than nonverbal subtest score (mean 75, range 53-98). Spatial ability was evaluated in a subset (n = 24) and was lower than verbal and nonverbal ability (mean 71, range 34-108). There was an average 14-point difference between verbal and nonverbal subset scores; 60% of the time the verbal subset score was higher than the nonverbal subset score. Study subjects with a verbal ability subtest score lower than the nonverbal subtest score were 4 times more likely to have a diagnosis of intellectual disability (suggestive, p-value 0.07). The age at which a child first spoke two-word phrases was strongly associated with measures of verbal ability (p-value 2.56e-07). Cognitive ability was correlated with adaptive behavior measures (correlation 0.42, p-value 0.02). However, though group means found equivalent score, there was, on average, a 10-point gap between these skills (range −33 to 33), in either direction, in about 50% of the sample suggesting that suggesting that cognitive measures only partially inform adaptive ability. Cognitive ability scores did not have any significant relationship to cumulative burden of psychopathology nor to individual neurodevelopmental or psychiatric diagnoses.

**Conclusions:** Individuals with 3q29 deletion syndrome have a complex pattern of cognitive disability. Two-thirds of individuals with the deletion will exhibit significant strength in verbal ability; this may mask deficits in non-verbal reasoning, leading to an over-estimation of overall ability. Deficits in verbal ability may be the driver of intellectual disability diagnosis. Cognitive ability is not a strong indicator of other neurodevelopmental or psychiatric impairment; thus individuals with 3q29 deletion syndrome who exhibit IQ scores within the normal range should receive all recommended behavioral evaluations.

## Introduction

Advances in research over the past 50-60 years have provided more fine-tuned approaches for describing the developmental differences of individuals with Intellectual and Developmental Disabilities (IDD). Originally, a more simplistic model of characterizing individuals with IDD highlighted primarily the cognitive deficits observed when compared to a typically developing standard (see (Zigler, 1969). This approach, however, failed to consider several factors.First, the imprecision of comparing individuals of the same chronological age without any reference for differences in developmental level.Second, the potential for etiological differences across individuals with IDD and thus unique phenotypic presentations. Lastly, the social and environmental factors influencing an individual’s cumulative life experience (For literature review see Zigler and Hodapp, 1986; Burack *et al*., 2001, 2012). Over time, an understanding of the methodological weaknesses to this approach emerged and a more inclusive manner of thinking about IDD began. This conceptual and methodological shift, focused on using a developmental approach to study what Zigler called the “whole child” (Zigler and Hodapp, 1986). Simply put, a way to more precisely describe an individual’s pattern of strengths and weaknesses across a variety of developmental domains (e.g., cognitive, social, adaptive) while also considering the various contexts (e.g., family, community, society) likely to impact the individual. Within this model, understanding cognitive skills remained important but were not the sole focus of characterizing individuals with IDD. Additionally, as technology in the field of genetics advanced, the ability to describe IDD based on known etiologic differences was further realized.

Given some of the extensive past problems of overgeneralizing findings of individuals with genetic syndromes secondary to lack of precision in defining participant groups to be studied, Zigler and his colleagues articulated the need of the developmental approach in understanding individuals with IDD (Zigler, 1967, 1969). Thus the study of a specific rare variant, in the case of this manuscript those with 3q29 deletion syndrome, can help us both understand the unique strengths and vulnerabilities of those individuals affected by the disorder as well as help our awareness of within- and between-group differences to better our understanding of the entire range of developmental problems, risk-potentiating factors, and protective characteristics (Burack, 1997). To help expand knowledge of IDD, we took lessons from the developmental formulation postulated by Zigler in the late 1960s, and the premise that “if the etiology of the phenotypic intelligence (as measured by an IQ) of two groups differs, it is far from logical to assert that the course of development is the same, or that even similar contents in their behaviors are mediated by exactly the same cognitive process” (Zigler, 1969, p. 553).

Here, we apply this knowledge to understand the cognitive profile of individuals with 3q29 deletion syndrome. 3q29 deletion syndrome (Hamosh, 2012) is caused by a hemizygous 1.6 Mb deletion containing 21 genes (Willatt *et al*., 2005; Ballif *et al*., 2008). The deletion is typically de novo, though inherited cases have been reported (Cox and Butler, 2015; Murphy *et al*., 2020). Early case reports describe mild to moderate intellectual disability as a common syndromic phenotype, with developmental delay, including speech delay, as the initial presentation (Girirajan *et al*., 2012; Cox and Butler, 2015). It is now understood that the 3q29 deletion is associated with a range of neurodevelopmental and psychiatric disabilities that manifest throughout the lifespan, including intellectual disability, autism, anxiety disorders, ADHD, and a 40-fold increased risk for schizophrenia (Mulle, 2015; Sanders *et al*., 2015; Glassford *et al*., 2016; Mulle *et al*., 2016; Marshall *et al*., 2017; Pollak *et al*., 2019; Sanchez Russo *et al*., 2021). In a recent study among individuals with the 3q29 deletion who were systematically evaluated with a deep-phenotyping protocol, only 34% qualified for a diagnosis of intellectual disability (Murphy *et al*., 2018; Sanchez Russo *et al*., 2021). IQ measures among study subjects ranged from 40-99, thus individuals with the 3q29 deletion may have cognitive ability that is well within the average range. This range of cognitive abilities tends to differ when intellectual functioning is described in case report studies which indicate a much greater percentage of individuals with intellectual disability. It also likely argues against prior reports estimating that 92% of individuals with the 3q29 deletion demonstrate a mild to moderate intellectual disability (Cox and Butler, 2015). These findings suggest the not only the possibility of an ascertainment bias, but may also conflate intellectual disability and learning disability, which are not one in the same. These biases may have had an outsized influence, overestimating the impact of the 3q29 deletion on cognitive ability.

The current study aims to carefully examine the cognitive profiles of individuals with 3q29 deletion syndrome, using cognitive data that has been uniformly measured among a case series of individuals with the 3q29 deletion ascertained from the 3q29 deletion registry (3q29deletion.org). Using Zigler’s developmental framework, we aim to identify the unique strengths and vulnerabilities of these individuals, as a group and individually, to better understand their developmental trajectory as well frame our findings onto other genetic disabilities or developmental disorders such as autism spectrum disorder.

## Methods

### Study Participants

Individuals with 3q29 deletion syndrome were recruited from the 3q29 registry (3q29deletion.org) housed at Emory University to participate in an in-person deep phenotyping study. The phenotyping protocol has been described previously (Murphy *et al*., 2018). Eligibility criteria were: a validated clinical diagnosis of 3q29 deletion syndrome where the subject’s deletion overlapped the canonical region (hg19, chr3:195725000-197350000) by ≥ 80%, and willingness and ability to travel to Atlanta, GA. Funding for travel was provided to increase the diversity of participants. Exclusion criteria were: any 3q29 deletion with less than 80% overlap with the canonical region; non-fluency in English, and age younger than six years. One exception to the age criterion was made; a 4.85-year-old who was part of a previously-described multiplex family was included in the current study (Murphy *et al*., 2020). Prior to study participation, an informed consent session was conducted, and repeated in-person at the beginning of the study visit. This study was approved by the Emory Institutional Board (IRB000088012).

### Instruments

Cognitive ability was evaluated using the Differential Ability Scales, Second Edition (DAS-II ages <18, n = 24) or the Wechsler Abbreviated Scale of Intelligence, Second Edition (WASI-II, >18, n = 7) which was determined based on the participants’ age. The DAS-II (Elliott, 2007) is an assessment of cognitive abilities for children ages 2 years, 6 months to 17 years, 11 months. The DAS-II is comprised of individual subtests that evaluate overall verbal, nonverbal and spatial abilities. A General Conceptual Ability (GCA) composite score is generated that reflects an overall estimate of cognitive functioning. The WASI-II (Wechsler, 2011) is an abbreviated measure of cognitive functioning for individuals ages 6-90 and provides an estimate of verbal and nonverbal abilities and generates an overall Full Scale IQ (FSIQ).

Of the individuals administered the DAS, 6 children aged 6 and younger were administered the DAS-II Early Years Battery (DAS-EY); one additional study subject, an 11 year old with limited verbal ability, was also administered the DAS-EY and age equivalents and ratio IQs were obtained. Seventeen children between ages 7-18 years were administered the DAS-II School Age Battery (DAS-SA). The other 8 individuals were administered the WASI-II. Instruments were administered by clinical psychologists (CS, CK, SW) early in the day to support the participants’ active engagement.

Adaptive behavior was assessed with the Vineland Adaptive Behavior Scales, Third Edition (Vineland-3; (Sparrow, Cicchetti and Saulnier, 2016). The Vineland-3 is a measure that assesses overall adaptive ability from birth to age 90 by gathering information about day-to-day activities across three adaptive domains of socialization, daily living skills, and communication skills. An Adaptive Behavior Composite (ABC) comprised of scores from each subdomain (Socialization, Daily Living Skills and Communication Skills) provides an overall estimate of an individual’s adaptive functioning. The Comprehensive Parent/Caregiver Form was utilized for this study. The Vineland-3 was completed by the parent or caretaker electronically via publisher websites (i.e., Pearson Q-global) either before or during the visit. Instruments were scored according to publisher’s instruction. Neurodevelopmental and psychiatric diagnoses were made using gold-standard instruments administered by experienced, trained professionals, as previously described (Sanchez Russo *et al*., 2021). Diagnoses were made by expert clinicians (CS, CK, SW, JB, LB, EW). To receive a diagnosis of ID, both cognitive and adaptive behavior scores needed to be approximately 2 or more standard deviations below the mean, consistent with diagnostic criteria in the Diagnostic and Statistical Manual of Mental Disorders, Fifth Edition (DSM-5; APA, 2013).

### Data Analysis

All data were captured in a custom local REDCap database. Data were exported for analysis and visualization in R. After confirming there were no test-specific score differences in standard scores for GCA/FSIQ, verbal ability (VIQ), and non-verbal ability (NVIQ) standard scores, data from the DAS-EY, DAS-SA, and WASI were combined across study subjects for all downstream analyses (Table S1, Figures S1-3).

## Results

### Participants

32 participants were consented and evaluated, including 4 participants from a single family. Study subjects ranged in age from 4.8-39.1 years (mean age 14.5 years, median age 11.7 years); 62.5% (n = 20) were male (Table 1).

### Profile of cognitive and adaptive abilities – overall group differences

Mean FSIQ across all study subjects was 73 (median 75.5, range 40-99). VIQ was slightly higher than FSIQ (mean = 80) and had greater variance across study subjects (median 85, range 31-106). NVIQ (mean 75, median 75, range 53-98) and spatial ability (mean 71, median 68, range 34-108) were both lower than verbal ability, though these differences were not statistically significant (Figure 1). Spatial ability was measured only with the DAS-II (n = 24 subjects) and was not analyzed further. There was a suggestive relationship between sex, for both FSIQ and VIQ, where female study subjects were on average 9 points higher for FSIQ (p-value 0.051) and 10 points higher for VIQ (p-value 0.08, Table S2). Because of the small sample size in this study (n = 12 females), we did not conduct any sex-stratified analyses, but we note that study of sex-related differences are an important future direction that will require a larger sample size. Age was not significantly associated with FSIQ, VIQ, or NVIQ (Table S3). The overall Adaptive Behavior Composite as measured by the Vineland-3 was a mean of 74 (median 71, range 48-110). Eleven individuals (34%) qualified for a diagnosis of intellectual disability (ID), where cognitive and adaptive behavior scores were ≥2 SD below the expected mean (≤ 70 on both evaluations).

**Figure 1.**
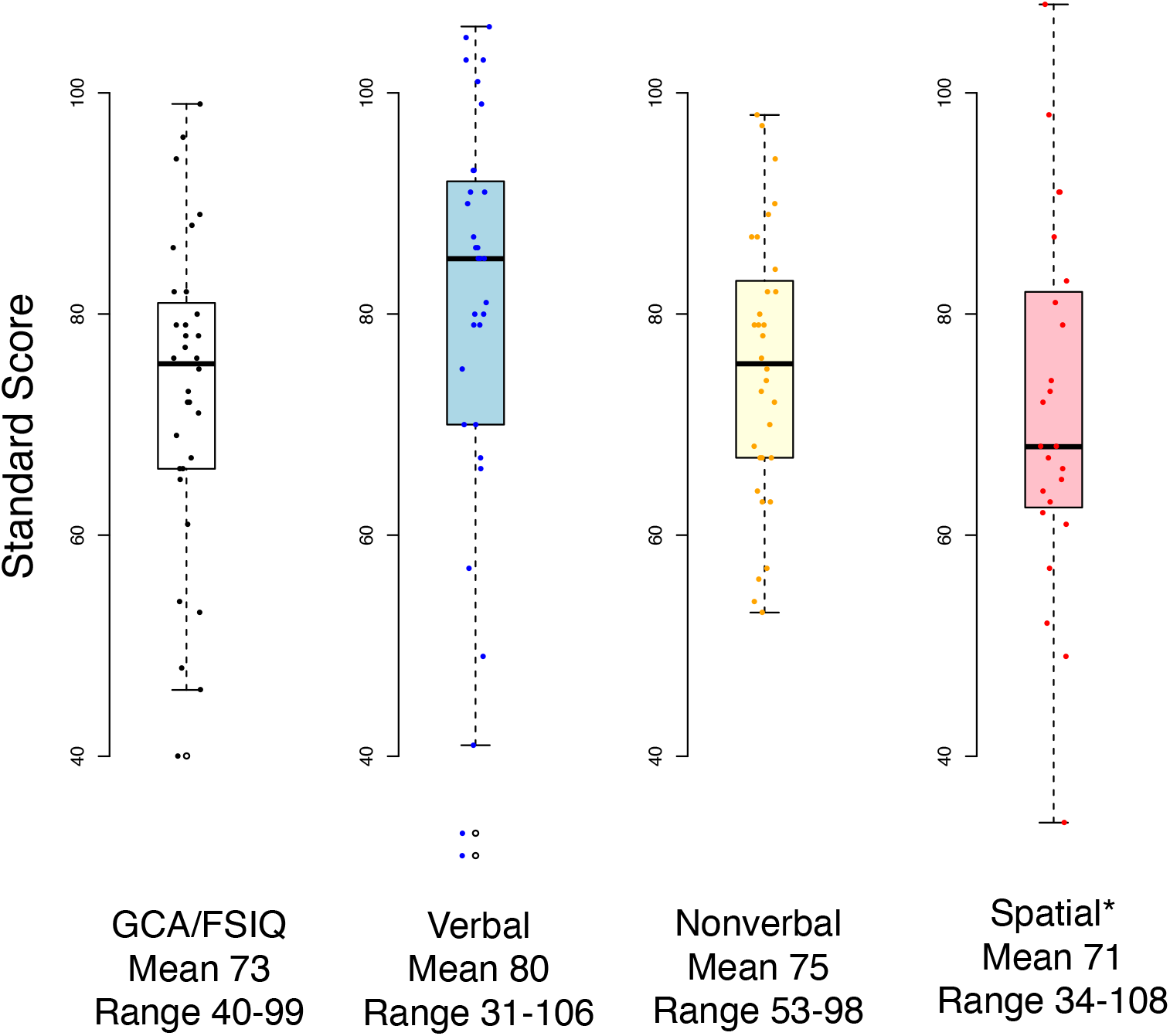
Cognitive ability measures in 3q29 deletion sydnrome.

### Relationship between cognitive ability and comorbid disorders

The 3q29 deletion is associated with a high burden of neurodevelopmental and psychiatric illness (Quintero-Rivera, Sharifi-Hannauer and Martinez-Agosto, 2010; Marshall *et al*., 2017; Pollak *et al*., 2019; Sanchez Russo *et al*., 2021). We examined whether FSIQ, VIQ, or NVIQ were associated with comorbid neurodevelopmental or psychiatric diagnoses common to our study subjects, including Autism Spectrum Disorder (ASD), Attention Deficit Hyperactivity Disorder (ADHD), any anxiety disorder, or psychosis. There was no relationship between FSIQ or VIQ and any individual diagnostic category (Tables S4, S5). For NVIQ, individuals with ASD had lower scores than those without ASD (69 vs 79, p = 0.02, Table S6). On the other hand, individuals with an anxiety disorder had higher NVIQ measures than those without an anxiety disorder (80 vs 72, p = 0.05, Table S6). These data suggest there may be complex interactions between cognitive ability and comorbid conditions, and a larger sample size will be required to untangle these relationships. We also sought to ask whether cognitive disability is related to the cumulative burden of neurodevelopmental and psychiatric illness. For each individual, we summed overall possible diagnoses (ASD, ADHD, anxiety disorder, psychosis). The total possible sum is 4; the range in our sample was 0-3 (0 diagnoses: 4 individuals; 1 diagnosis, 13 individuals; 2 diagnoses, 9 individuals; 3 diagnoses, 6 individuals). Our data (Figure 2) do not suggest any relationship between cognitive ability and overall burden of illness (p-value 0.92); individuals with 3q29 deletion syndrome who escape a substantial cognitive impact (with cognitive ability scores in the normal range) remain at significant risk for additional neurodevelopmental and psychiatric comorbidity.

**Figure 2.**
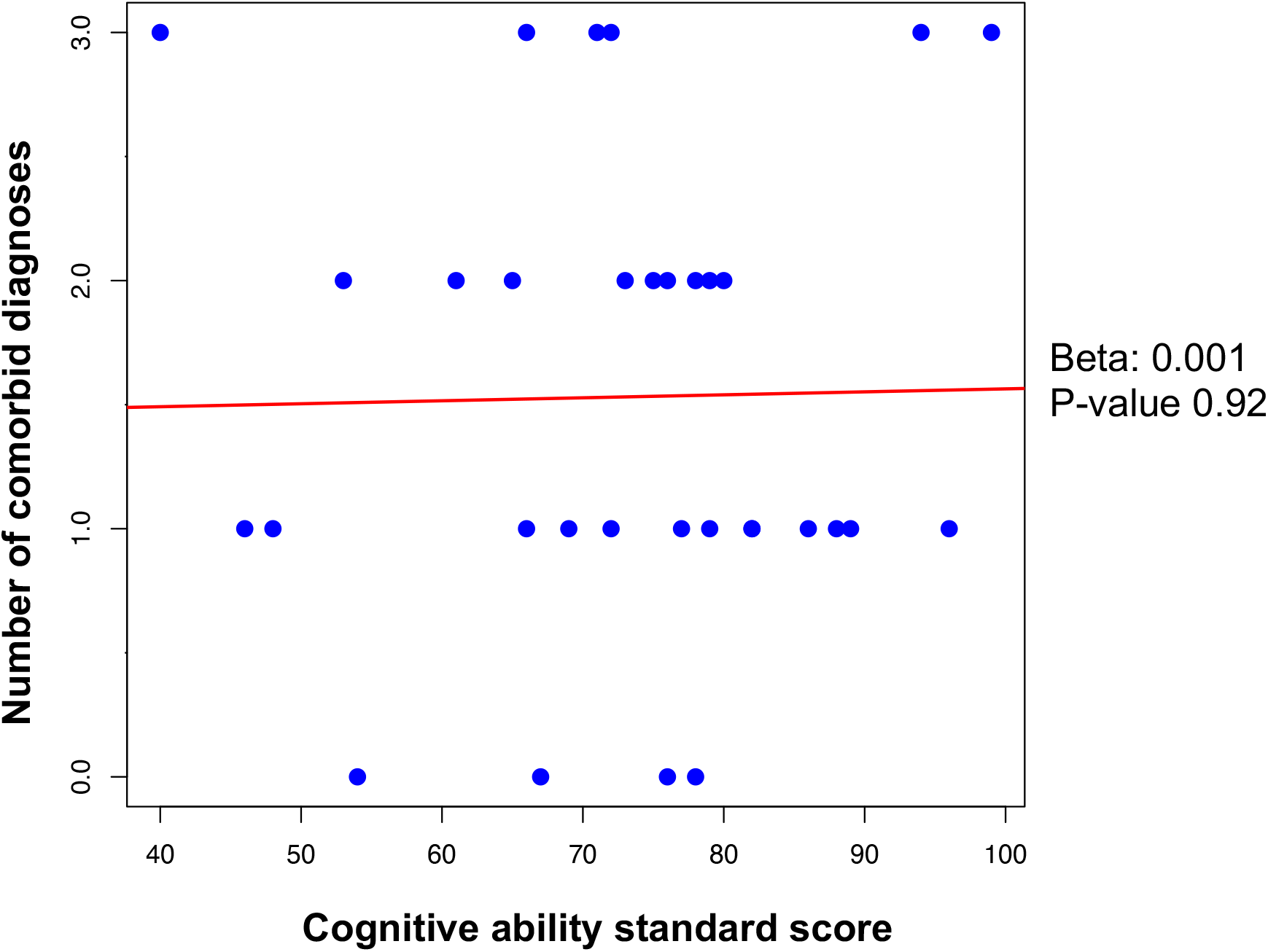
Relationship between cognitive ability and comorbid diagnoses.

### Differences in IQ subtest scores in 3q29 deletion syndrome – individual subject results

In the majority of our study subjects, at an individual data level, there were large discrepancies between verbal and nonverbal subtest scores (Figure 3A and 3B). The average absolute difference between subtest scores was 14 points. In 59% of our sample (n = 19), the verbal subtest score was 5 or more points higher than the nonverbal score (range 5-42). For 31% of our sample (n = 10), the verbal subtest score was 5 or more points lower than the nonverbal subtest score (range 7-34). For 9% of our samples (n = 3), verbal and nonverbal subset scores were within 5 points of one another.

**Figure 3.**
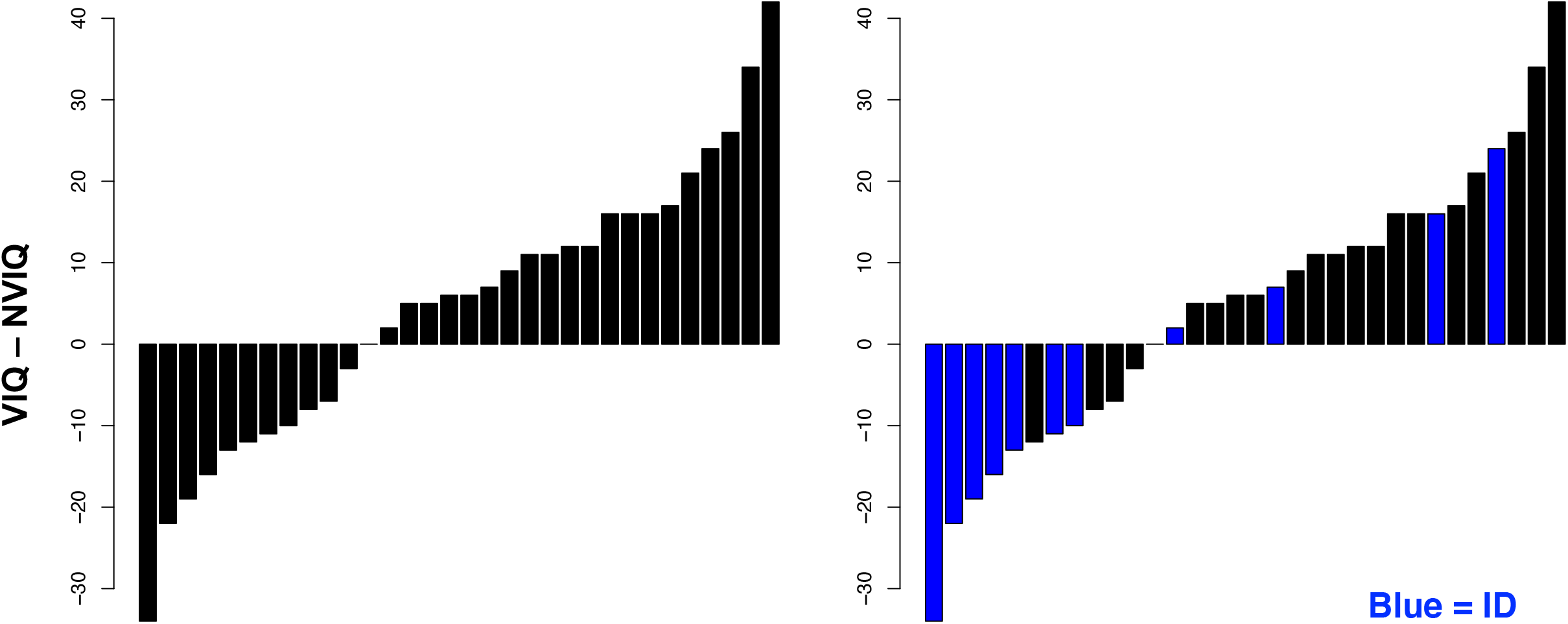
A pronounced VIQ/NVIQ split in 3q29 deletion syndrome. Low VIQ may be associated with ID.

We examined the relationship between VIQ/NVIQ disparity and comorbid diagnoses. Of 10 individuals with VIQ lower than NVIQ, 7 (70%) had a diagnosis of ID, compared to 4 out of 22 individuals (18%) with VIQ higher than or equal to NVIQ (odds ratio 9.5, p-value 0.01). This analysis reveals that among individuals with 3q29 deletion syndrome, those with VIQ lower than NVIQ are 9.5 times more likely to have a diagnosis of intellectual disability, as compared to individuals with VIQ equal to or higher than NVIQ. There were no significant relationships between the VIQ/NVIQ difference and other comorbid phenotypes (Figures S4A-D).

### Predictors of verbal ability

Verbal ability was a noted strength for many of our study subjects. This is a slight paradox, as many individuals with 3q29 deletion syndrome exhibit speech delay. In our study sample, the average age that children began saying first words was 1.6 years (range 0.5 – 4 years), and the average age for speaking in two-word phrases was 3.2 years (range 1.2-10 years). We evaluated whether verbal ability was “in process” and still developing for our study subjects, by assessing whether the time in years from talking (first words or two-words phrases) to the time of our evaluation was associated with verbal ability. There was no apparent relationship between these variables, revealing that the VIQ-NVIQ disparity was not an artifact of developmental time (Figure S5). We next evaluated whether absolute age at talking (single words or two-word phrases) was associated with verbal ability. Age at which the child spoke two-word phrases was strongly associated with verbal ability (p-value < 0.0001) with an estimated decrease of 8 points in VIQ for every year that speaking in two-word phrases was delayed. This relationship was specific to two-word phrases; the age at which single words were spoken was not associated with VIQ (p-value 0.93). Speaking in two words-phrases had at best a suggestive relationship to NVIQ (p-value 0.06), with an estimated loss of 2 NVIQ points for every year two-word phrases were delayed.

### Relationship between cognitive ability and adaptive behavior

FSIQ and adaptive behavior standard scores were well-correlated (correlation coefficient 0.42, p-value 0.02, Figure 5). However, 50% of our study sample had a departure of half a standard deviation (7.5 points) or more (average mean absolute difference between cognitive and adaptive standard scores = 10.9). Six study subjects (19%) had lower adaptive behavior scores than would be expected based on their cognitive ability; 10 study subjects (31%) had higher adaptive behavior scores than would be expected based on cognitive ability. There was no relationship between comorbid diagnosis and cognitive/adaptive disparity. VIQ standard scores were on average 5 points lower than adaptive behavior standard scores (range 27 to −38, absolute mean difference 13 points, Figure 5), and many study subjects with high verbal ability perform lower than expected for adaptive behaviors. NVIQ was also likely to depart from adaptive behavior standard scores, but was less likely to systematically be lower (NVIQ on average 1 point lower than adaptive behavior standard scores, range 29 to −30, absolute mean difference 11 points).

**Figure 4.**
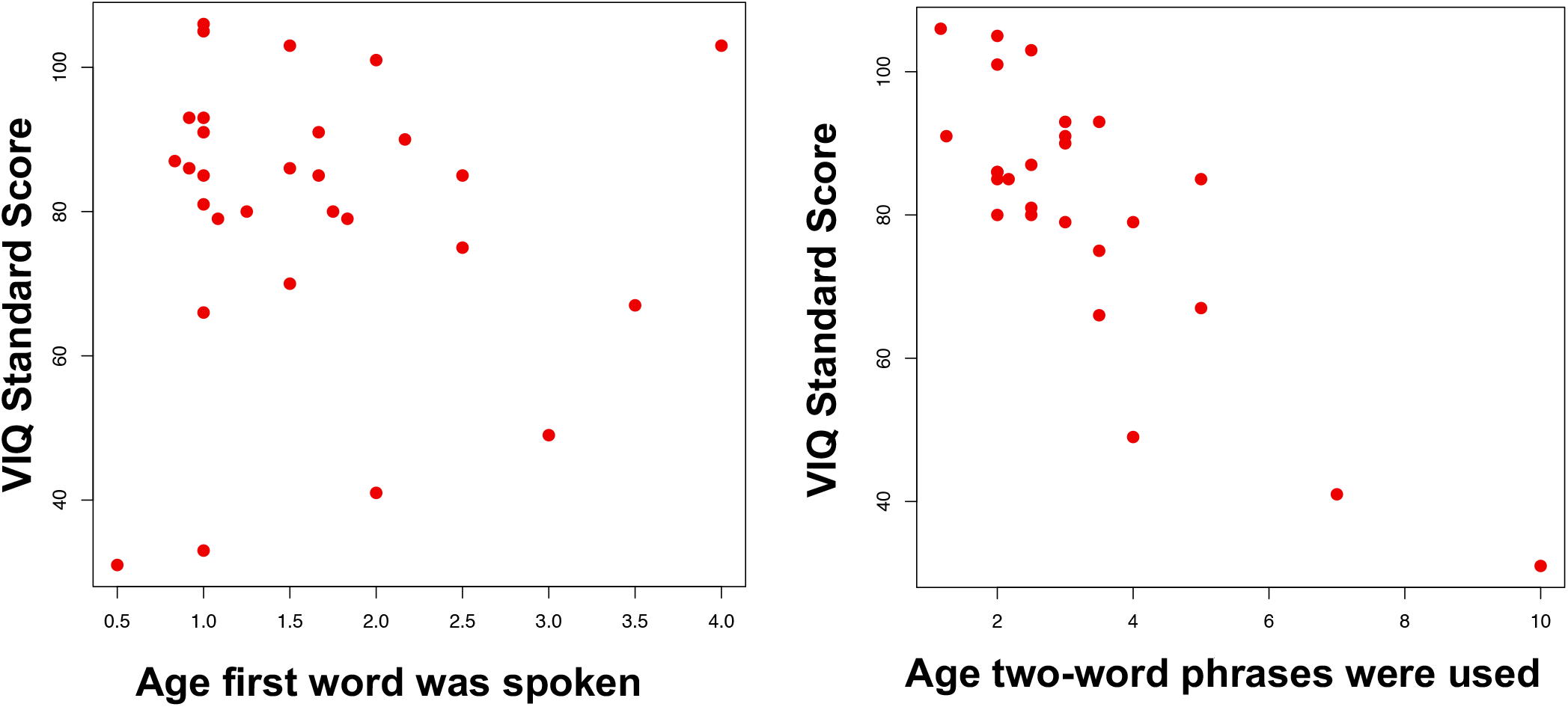
Age at first two-word phrase (but not first word) is highly correlated with VIQ.

**Figure 5.**
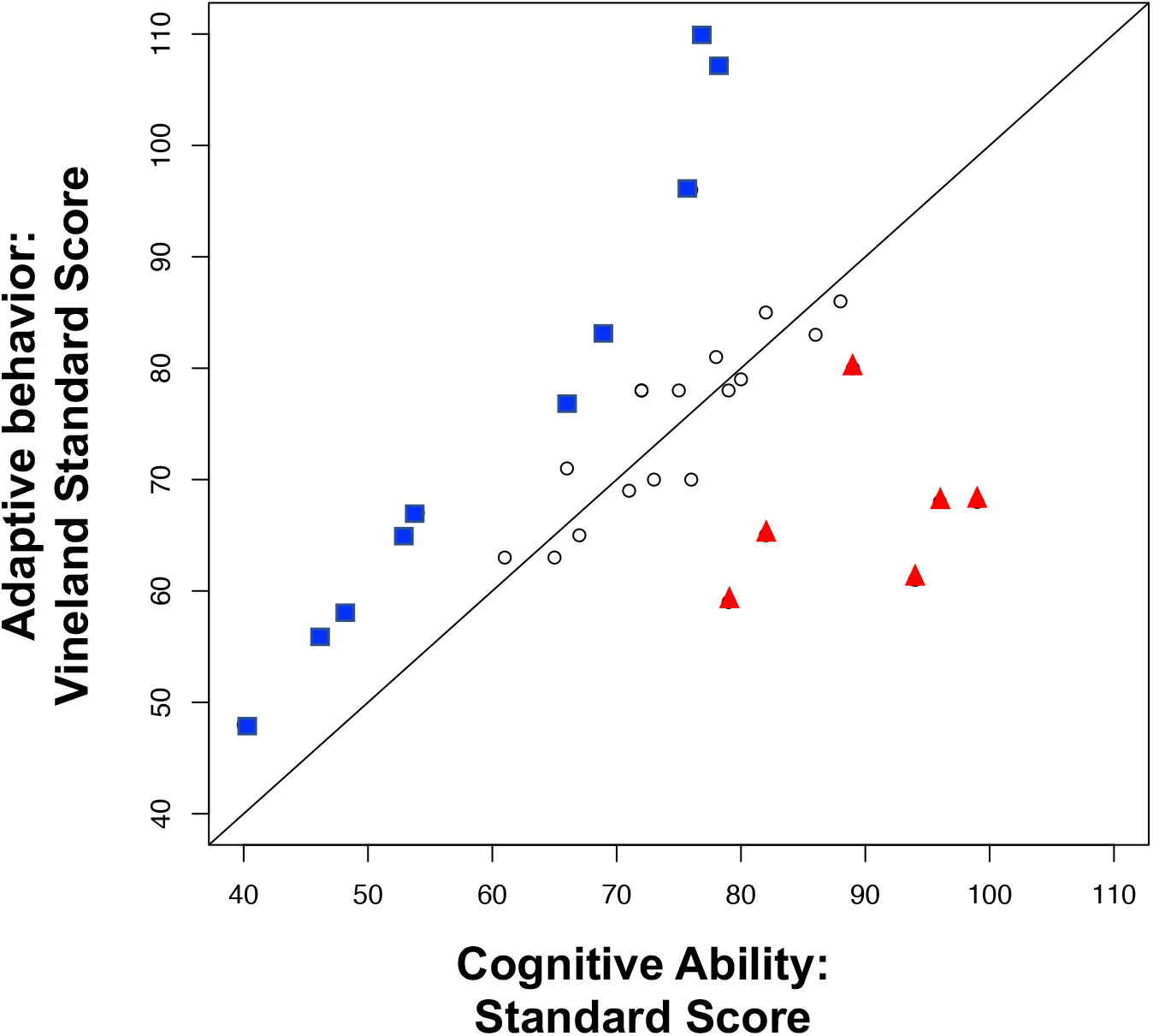
Cognitive ability is poorly predictive of adaptive behavior.

## Discussion

### Overall Cognitive Profile

Individuals with 3q29 deletion syndrome demonstrate a unique profile of cognitive strengths and weaknesses. While the average overall IQ across participants was in the range of what would be considered mild intellectual disability, there was variability across individual subjects ranging from moderate intellectual disability to average cognitive functioning. This finding supports the variability observed in case reports, particularly when the 3q29 deletion is inherited (Li *et al*., 2009; Cobb *et al*., 2010; Khan *et al*., 2019; Murphy *et al*., 2020). Additionally, in line with the developmental perspective, looking at differences across verbal, non-verbal, and spatial skills reveals that this overall IQ may be less meaningful in understanding the functioning level of individuals with 3q29 deletion syndrome as there are specific strengths and weaknesses noted in these individual domains. For example, verbal ability tended to be an area of relative strength across participants whereas nonverbal and spatial skills (where collected) were areas of relative weakness. This pattern of strengths and weaknesses may set the stage for possible learning difficulties or disabilities in this group. Mild to moderate learning disabilities were similarly reported in an estimated 92% of individuals with the 3q29 deletion syndrome (Cox and Butler, 2015). However, learning disabilities are often diagnosed in context of academic difficulties and so these assumptions are provided tentatively and as an area that requires further investigation. Although anecdotally many of the children in the study did require academic supports through their local school district, receiving some level of special education services.

With regard to specific cognitive profiles, verbal ability was a noted strength for many of our study subjects despite the reported history of speech delay highlighting the difference between communication skills and verbal cognitive abilities. With each year of delay in speaking in phrases, individuals with 3q29 had an estimated decrease of 8 points in their verbal cognitive abilities. No other developmental factor or comorbid psychiatric or developmental disability was related to cognitive abilities or splits between verbal and nonverbal abilities. In addition, there was no relation between age, sex or age at first words indicating that the verbal/nonverbal cognitive disparity was not an artifact of developmental time.

### Cognitive Functioning and Adaptive behavior

In our sample, 11 individuals met criteria for some level of Intellectual Disability demonstrating scores on both cognitive and adaptive measures significantly below the average range (i.e., ≥2 SD below the expected mean). This is a 31-fold increase over the expected population prevalence of 1.1% (Zablotsky *et al*., 2019).Although overall cognition and adaptive behavior scores were well-correlated in our sample, individual profiles of cognition and adaptive behavior varied significantly across participants with 3q29 deletion syndrome. While some individuals skills were consistent across measures, 50% of the sample presented with adaptive skills that were either slightly higher than expected (31%) or lower than expected. Furthermore, many individuals with higher overall verbal skills on cognitive assessments performed lower on adaptive measures. Given that verbal ability is readily apparent and easily observed in many settings, there is a risk that overall ability may be overestimated for these individuals. These data underscore that mean scores and scores presented in isolation (i.e. just one construct) are limited in informing about overall profiles and outcomes for individuals with 3q29 deletion syndrome.

### Cognitive Functioning and Comorbid Disorders

The 3q29 deletion is associated with a high burden of comorbid neurodevelopmental and psychiatric diagnoses. In our study sample, there was no association between overall cognitive abilities or verbal IQ and the assessed diagnostic categories of Autism Spectrum Disorder (ASD), Attention Deficit Hyperactivity Disorder (ADHD), any anxiety disorder, or psychosis. Individuals with ASD had lower nonverbal IQ scores than those without ASD, and individuals with anxiety disorder had higher nonverbal IQ than those without an anxiety disorder. These data suggest there may be complex interactions between cognitive ability and comorbid conditions, and a larger sample size will be required to untangle these relationships. We also sought to ask whether cognition is related to the cumulative burden of neurodevelopmental and psychiatric conditions. Overall, there was no relationship between intellectual disability and the overall burden of co-occurring psychiatric conditions in individuals with 3q29 deletion syndrome. However, individuals without cognitive delay remain at significant risk for having co-occurring neurodevelopmental and psychiatric conditions. Thus, individuals with 3q29 deletion syndrome with and without cognitive impairment should receive all recommended behavioral evaluations.

### Summary

These findings underscore the importance of taking a developmental approach in the assessment and characterization of individuals with 3q29 deletion syndrome. Individuals with 3q29 deletion syndrome do not show a simple phenotypic pattern but instead are a complex interplay of cognitive and adaptive profiles, language development, and co-occurring anxiety, ASD, ADHD, and psychosis. Overall IQ is not the best predictor of outcomes in these individuals, particularly given the cognitive splits in the majority of individuals. These gaps in cognitive skills may also be an indicator of the reported rates of learning disabilities and challenges faced in an academic setting.

Limitations of this study include the relatively small sample size, particularly when examining cognitive profiles in psychiatric comorbidities. That being said, this is the largest sample of individuals with 3q29 deletion syndrome assessed to date. In addition, despite funding for travel provided by the study, the sample includes those individuals who were willing and able to travel to Atlanta. This may represent ascertainment bias; future studies are needed to measure IQ and psychiatric comorbidities in a virtual manner or with a study design where phenotypic experts can travel to the homes of participants rather than requiring individuals to travel to Atlanta. Additional studies will also explore the impact of executive functioning on cognitive profiles as well as profiles of academic functioning.

## Collaborators

Emory 3q29 Project: <u>Hallie Averbach, Gary J Bassell, Shanthi Cambala, Tamara Caspary, David Cutler, Paul A Dawson, Michael P Epstein, Henry R Johnston, Bryan Mak, Tamika Malone, Trenell Mosley, Ava Papetti, Rebecca M Pollak, Ryan Purcell, Nikisha Sisodoya, Steven Sloan, Stephen T Warren, David Weinshenker, Zhexing Wen, Mike Zwick</u>

## Data Availability

Data are available to qualified investigators upon reasonable request through the NIMH Data Archive.

## Acknowledgements

We acknowledge the 3q29 study subjects and their families. We also acknowledge The Emory 3q29 Project members. We gratefully acknowledge The Marcus Autism Center for providing clinical assessment resources. Funding for this work was provided by NIH grants R01 MH110701 and R01 MH118534. REDCap is supported by grant UL1 TR000424.

## Ethics Declaration

Informed consent was received from all participating study subjects. This study was approved by the Emory Institutional Board (IRB000088012).

