## Supplementary material for "A distinct cognitive profile in individuals with 3q29 deletion syndrome": Figure legends

**Figure 1:** Box plots and distribution of scores for overall cognitive ability, verbal ability, nonverbal ability, and spatial ability for 32 study subjects with 3q29 deletion syndrome. *Spatial ability measured in a subset of 24 individuals who were evaluated with the DAS.

**Figure 2:** The relationship between cognitive ability standard score and cumulative number of comorbid diagnoses is shown. Possible diagnoses included ASD, ADHD, anxiety disorders, and psychosis. Total possible number of comorbid diagnoses was 4, the range was 0-3. Individuals with higher IQ scores have an equivalent burden of comorbid neurodevelopmental and psychiatric diagnoses when compared to individuals with lower IQ scores.

**Figure 3:** Panel A: for each individual, the difference between their verbal and nonverbal IQ scores are plotted. On average, verbal and nonverbal scores are 14 points apart, in either direction. 10 individuals (31% of the sample) have verbal IQ scores that are more than 5 points lower than nonverbal scores. 19 individuals (59%) have verbal IQ scores that are at least 5 points higher than nonverbal scores. Only three individuals in this study (9%) have VIQ and NVIQ scores within 5 points of each other. Panel B: The same plot, showing individuals with a diagnosis of ID in blue. 7 out of 10 (70%) individuals with lower VIQ then NVIQ have an ID diagnosis, compared to 4 of 22 (18%) individuals where VIQ is equal to or higher than NVIQ (p-value 0.01). This 9.5-fold enrichment suggests that for individuals with 3q92 deletion syndrome, VIQ lower than NVIQ is a risk factor for ID.

**Figure 4:** Panel A: Age at which the first word is spoken (as evaluated by the ADI-R) is not associated with verbal ability (p-value 0.92). Panel B: Age at which two-word phrases is strongly associated with verbal ability (p-value 0.0000003). The linear regression beta estimate of -8 reveals that for every year a child is delayed speaking in two word phrases, verbal ability scores are projected to be 8 points lower.

**Figure 5:** Cognitive ability is only partially predictive of adaptive behavior. Blue squares (n = 10) show individuals with 3q29 deletion syndrome who perform at least 7.5 points higher on adaptive behavior measures than would be predicted by cognitive ability; red triangles (n = 6) show individuals with 3q29 deletion syndrome who perform at least 7.5 points lower on adaptive behavior measures than would be predicted by cognitive ability. Only 16 individuals (50%) perform as expected on adaptive behaviors measures, based on cognitive ability scores.
