## supplemental files for "A distinct cognitive profile in individuals with 3q29 deletion syndrome"

Supplemental data:

1. Comparison of cognitive test and subtest scores across instruments
2. Relationship between sex and age to cognitive test and subtest scores
3. Relationship between test and subset scores and diagnosis
4. Relationship of verbal/nonverbal score split to diagnosis
5. Time in years between age at talking and study assessment: relationship to observed verbal ability

1. Comparison of cognitive test and subtest scores across instruments:

In this study, we used three instruments to assess cognitive ability, depending on the age and developmental level of the test subject: Differential Ability Scales-Second Edition, Early Years (DAS-EY, n = 6); Differential Ability Scales, School Age (DAS-SA, n = 17); and the Weschler Abbreviated Intelligence Scales-Second Edition (WASI, n = 8). To assess whether test and subtest scores could be combined across tests, we visualized the data and performed statistical tests to confirm the scores were drawn from approximately the same distributions. These data reveal there may be suggestive evidence for test-related differences in non-verbal ability that this study is underpowered to detect. A larger sample size is required to determine if these are true differences. Given the global similarity in scores across tests, we chose to combine them for downstream analyses.

**Table S1: Comparison across instruments**

| Comparison | FSIQ/GCA | Verbal Ability | Non-verbal ability |
| --- | --- | --- | --- |
| DAS-EY vs DAS-SA | Mean DAS EY: 74  Mean DAS-SA: 72  p-value: 0.79 | Mean DAS-EY: 76  Mean DAS-SA: 79  p-value: 0.74 | Mean DAS-EY: 84  Mean DAS-SA:73  p-value: 0.06 |
| DAS-EY vs WASI | Mean DAS-EY: 74  Mean WASI: 76  p-value: 0.83 | Mean DAS-EY: 76  Mean WASI: 83  p-value: 0.51 | Mean DAS-EY: 84  Mean WASI: 72  p-value: 0.12 |
| DAS-SA vs WASI | Mean DAS-SA: 72  Mean WASI: 76  p-value: 0.44 | Mean DAS-SA: 79  Mean WASI: 83  p-value: 0.64 | Mean DAS-SA: 73  Mean WASI: 72  p-value: 0.84 |

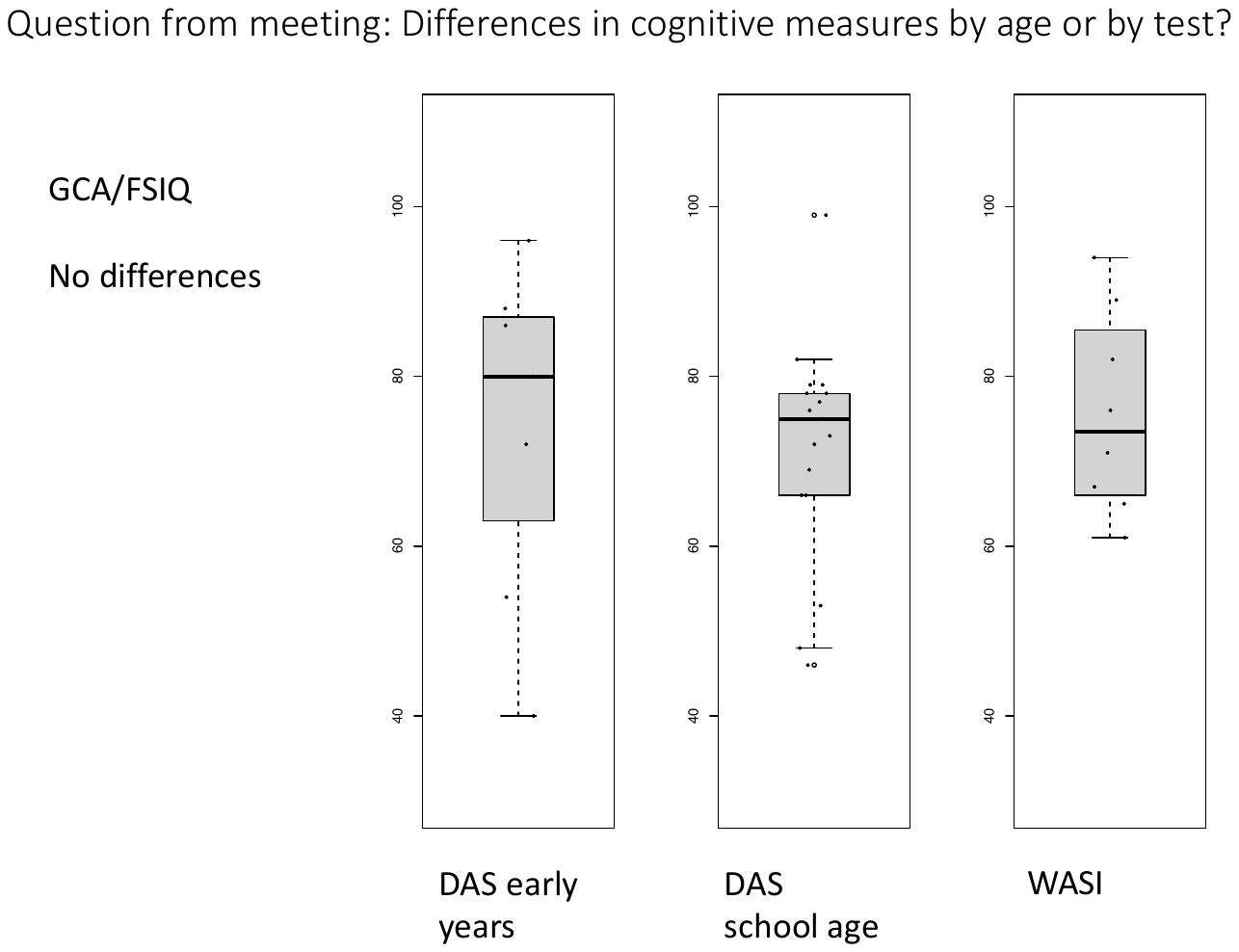

**Figure S1:** No differences in GCA/FSIQ between instruments.

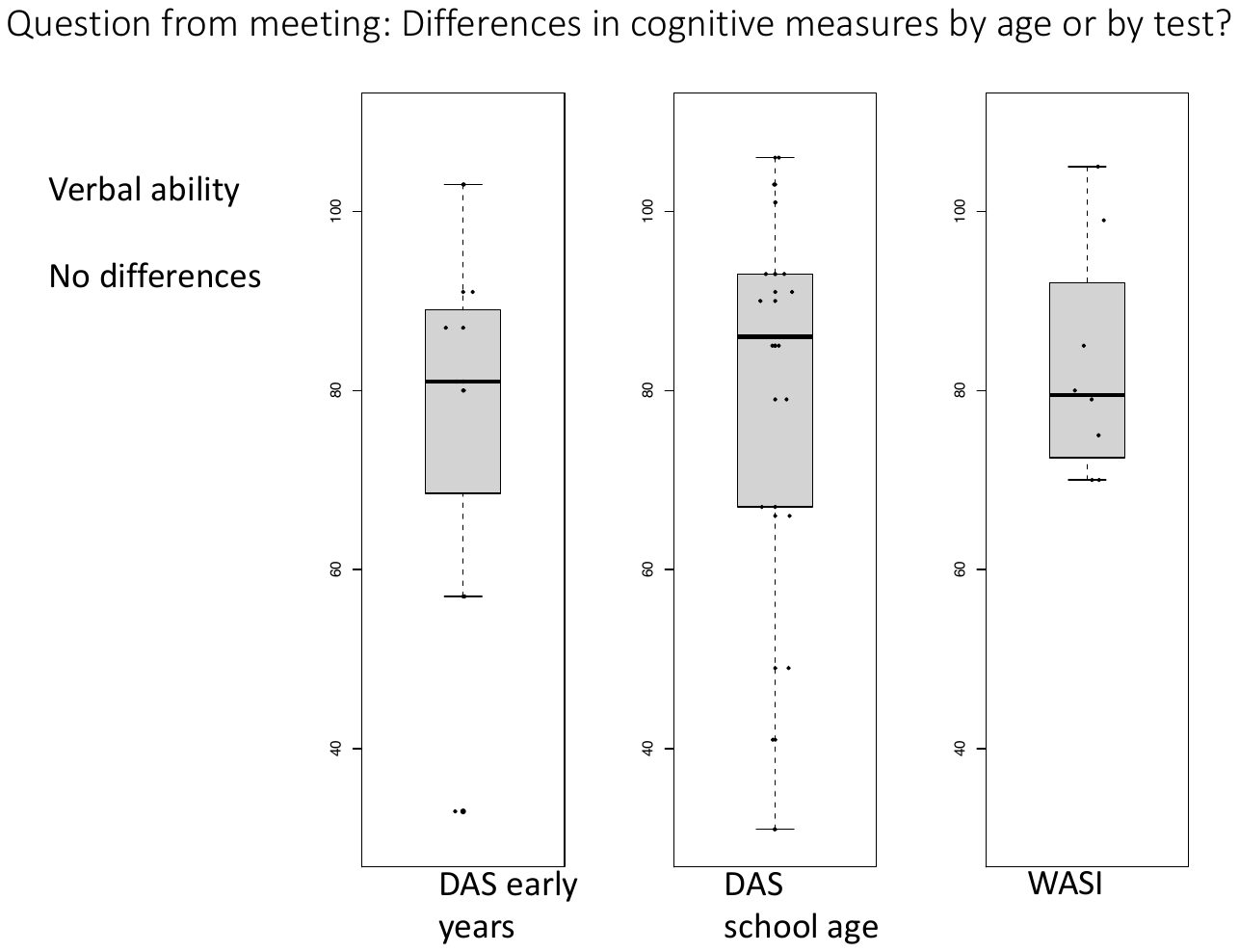

**Figure S2:** No differences in verbal ability between instruments.

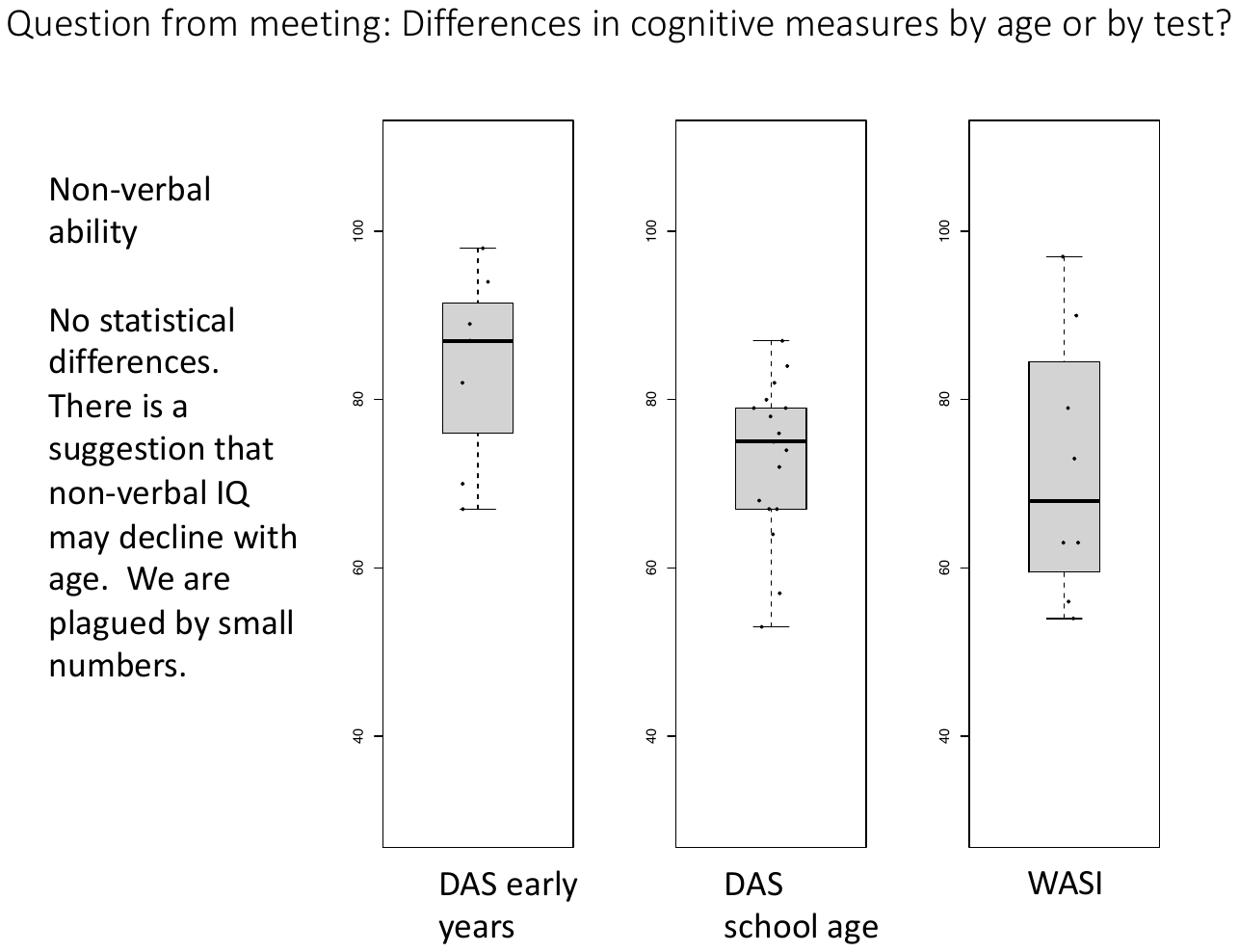

**Figure S3:** Suggestive differences in nonverbal ability between instruments, with the DAS-EY showing higher measures than other instruments.

2. Relationship of sex and age to cognitive test and subtest scores

After data were combined across instruments, we sought to determine whether there were any sex- or age-dependent relationships between IQ scores or subtest scores in our sample. To do this, we performed t-tests (for sex) or linear regression (for age and age + sex). There was a suggestive relationship between sex, for both FSIQ and VIQ, where female study subjects were on average 9 points higher for FSIQ (p-value 0.051) and 10 points higher for VIQ (p-value 0.08). Because of the small sample size in this study (n = 12 females), we did not conduct any sex-stratified analyses, but we note that study of sex-related differences are an important future direction that will require a larger sample size. There were no significant relationships between age and cognitive variables in our study.

**Table S2: Cognitive test and subtest scores by Sex**

|  | Overall cognitive ability | Verbal ability | Non-verbal ability |
| --- | --- | --- | --- |
| Male mean | 70 | 76 | 73 |
| Female mean | 79 | 86 | 78 |
| p-value | 0.051 | 0.08 | 0.23 |

**Table S3: Cognitive test and subset scores by age and age + sex:**

| **Model** | **Beta Estimate(s)** | **P-value** |
| --- | --- | --- |
| FSIQ by age | Age: 0.14 | 0.6 |
| FSIQ by age, sex | Age: 0.08 | 0.8 |
|  | Sex: 9.0 | 0.09 |
| Verbal IQ by age | Age: 0.3 | 0.5 |
| Verbal IQ by age, sex | Age: 0.2 | 0.6 |
|  | Sex: 10.4 | 0.2 |
| NVIQ by age | Age: -0.4 | 0.1 |
| NVIQ by age, sex | Age: -0.4 | 0.1 |
|  | Sex: 6 | 0.2 |

3. Relationship between cognitive test and subset scores and diagnosis

We next asked whether there were any relationships between cognitive ability scores or subtest scores and any of the neurodevelopmental or psychiatric diagnoses common to our study subjects. We saw no relationships with FSIQ:

**Table S4: Cognitive test scores and comorbid diagnosis**

| **DX** | **Yes: FSIQ mean** | **No: FSIQ mean** | **P-value** |
| --- | --- | --- | --- |
| ASD | 68 | 76 | 0.12 |
| ADHD | 75 | 70 | 0.31 |
| Any anxiety disorder | 76 | 71 | 0.39 |
| psychosis | 73 | 74 | 0.88 |
| ID | 59 | 81 | 0.00002 |

There were also no relationships between diagnosis and verbal IQ measures:

**Table S5: VIQ test scores and comorbid diagnosis:**

| **DX** | **Yes: VIQ mean** | **No: VIQ mean** | **P-value** |
| --- | --- | --- | --- |
| ASD | 72 | 84 | 0.16 |
| ADHD | 80 | 80 | 0.99 |
| Any anxiety disorder | 81 | 79 | 0.69 |
| psychosis | 83 | 82 | 0.94 |
| ID | 58 | 91 | 0.00005 |

Suggestive relationships between diagnosis and NVIQ were identified. Individuals with ASD scored on average 10 points *lower* on NVIQ measures. Individuals with anxiety disorders scored on average 8 points *higher* for NVIQ measures. These data suggest there may be complex interactions between cognitive ability and comorbid conditions, and a larger sample size will be required to untangle these relationships.

**Table S6: NVIQ test scores and comorbid diagnosis:**

| **DX** | **Yes: NVIQ mean** | **No: NVIQ mean** | **P-value** |
| --- | --- | --- | --- |
| ASD | 69 | 79 | 0.02* |
| ADHD | 78 | 71 | 0.15 |
| Any anxiety disorder | 80 | 72 | 0.05* |
| psychosis | 69 | 75 | 0.49 |
| ID | 65 | 80 | 0.0002 |

4. Relationship of verbal/nonverbal score split to diagnosis

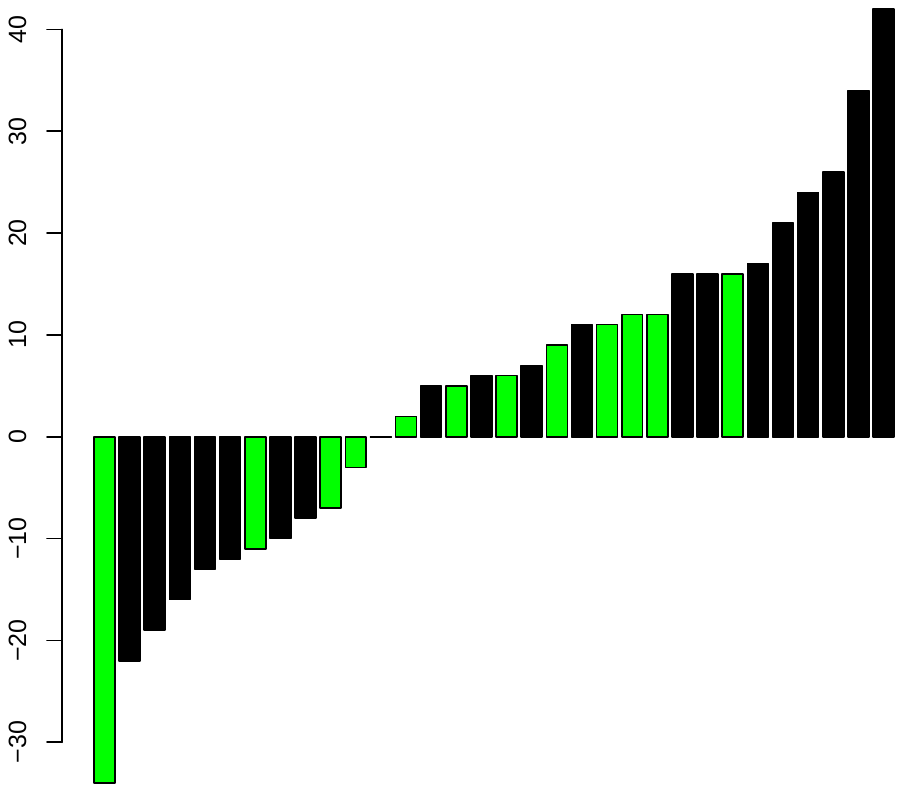

**Figure S4C:** Y-axis, verbal-nonverbal ability for 32 study subjects with 3q29 deletion. Purple bars indicate anxiety disorder diagnosis.

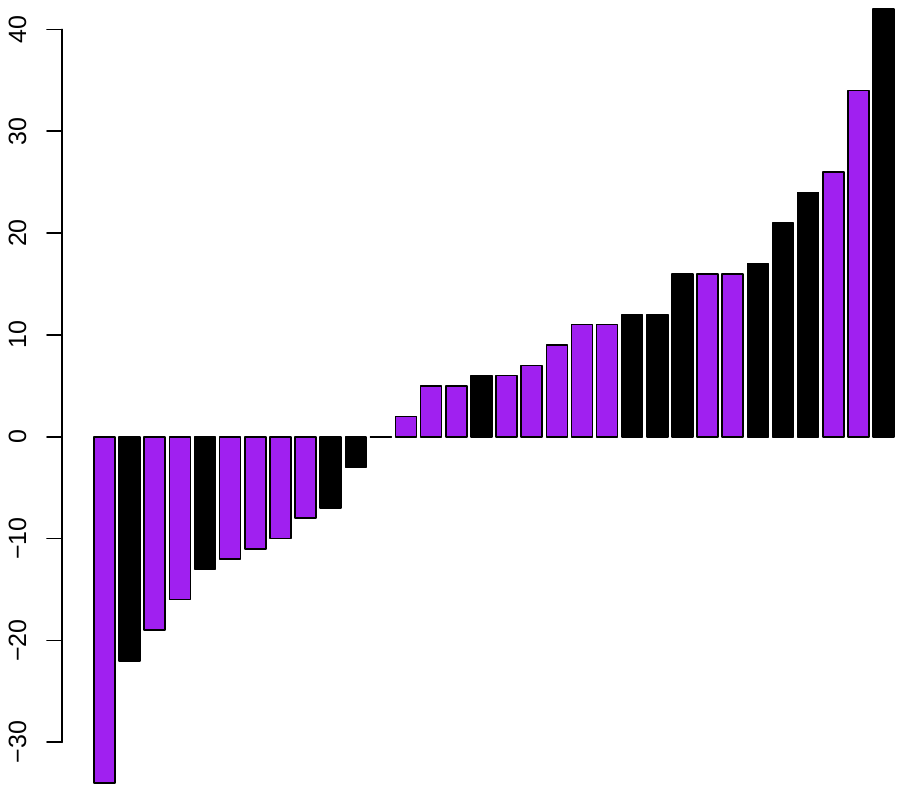

**Figure S4A**: Y-axis, verbal-nonverbal ability for 32 study subjects with 3q29 deletion. Purple bars indicate ADHD diagnosis.

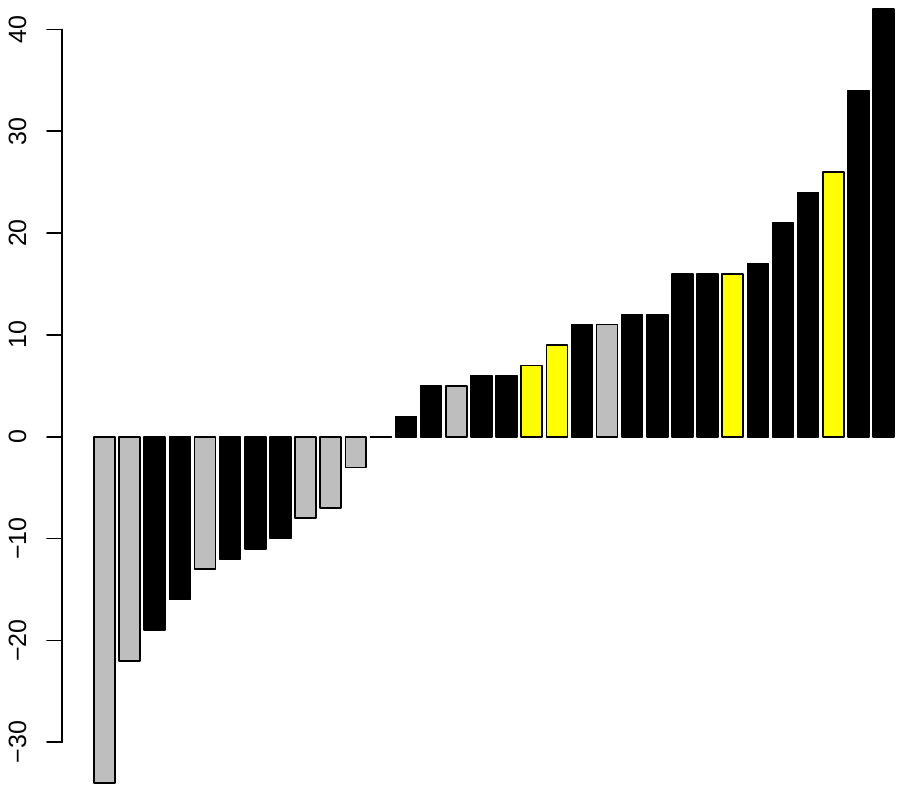

**Figure** **S4D**: Y-axis, verbal-nonverbal ability for 32 study subjects with 3q29 deletion. Yellow bars indicate diagnosis of psychosis (gray bars = individual not assessed for psychosis).

**Verbal – nonverbal subtest scores**

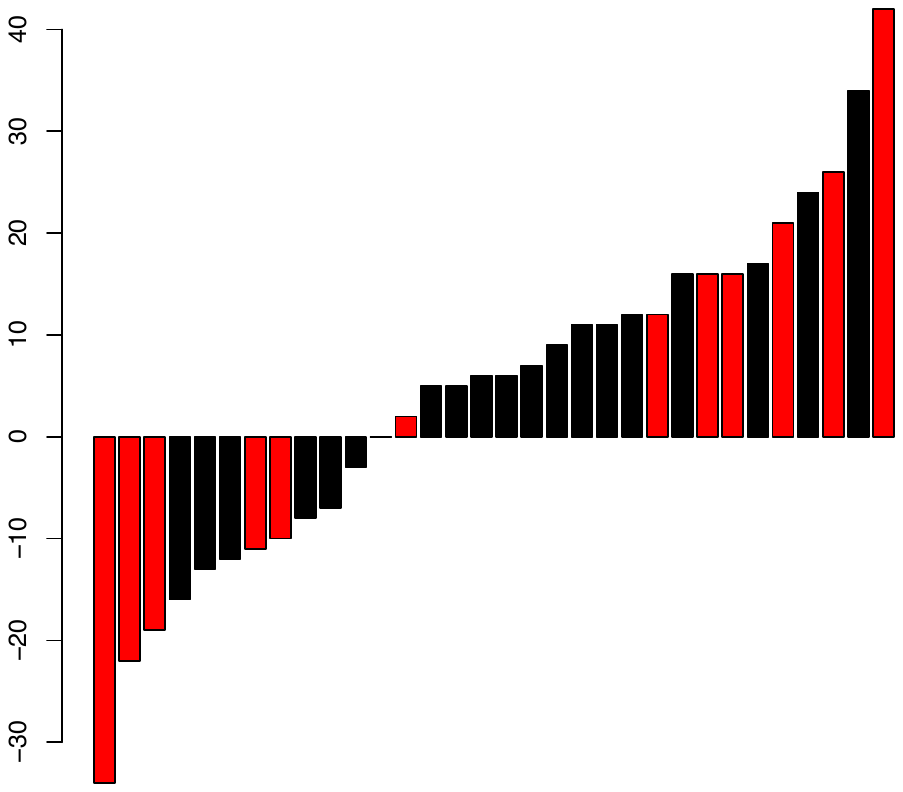

**Figure S4B:** Y-axis, verbal-nonverbal ability for 32 study subjects with 3q29 deletion. Red bars indicate ASD diagnosis.

We sought to identify whether the pronounced verbal/nonverbal score split was related in any way to other comorbid neurodevelopmental or psychiatric diagnoses present in our study subjects. Aside from the suggestive relationship to intellectual disability discussed in the main text of the paper, no relationships were identified.

5. Time in years between age at talking and study assessment: relationship to observed verbal ability

To assess whether aspects of our study design were creating artifactual relationships to verbal ability scores, we assessed whether the time between when a child started talking and the age at our assessment was related to their verbal ability score. To put it another way: If one child began talking at age 2 and was assessed at age 7, that child would have had five years to develop their verbal ability. Another child who began talking at age 4 and was assessed at age 7, would have had only three years to develop their verbal ability, and might be predicted to have lower verbal ability scores. To assess this relationship, we extracted two variables from the Autism Diagnostic Interview-Revised (ADI-R),which was applied uniformly to all individuals in our study. The variables we extracted were “age first word was spoken” and “age two-word phrases were spoken.” We evaluated whether the time in years between age at first word/age at two word phrases and our evaluation was associated with verbal ability scores. Our data revealed no significant relationship between these variables (using a simple linear regression framework). This analysis shows that our study design did not have undue influence on verbal ability scores.

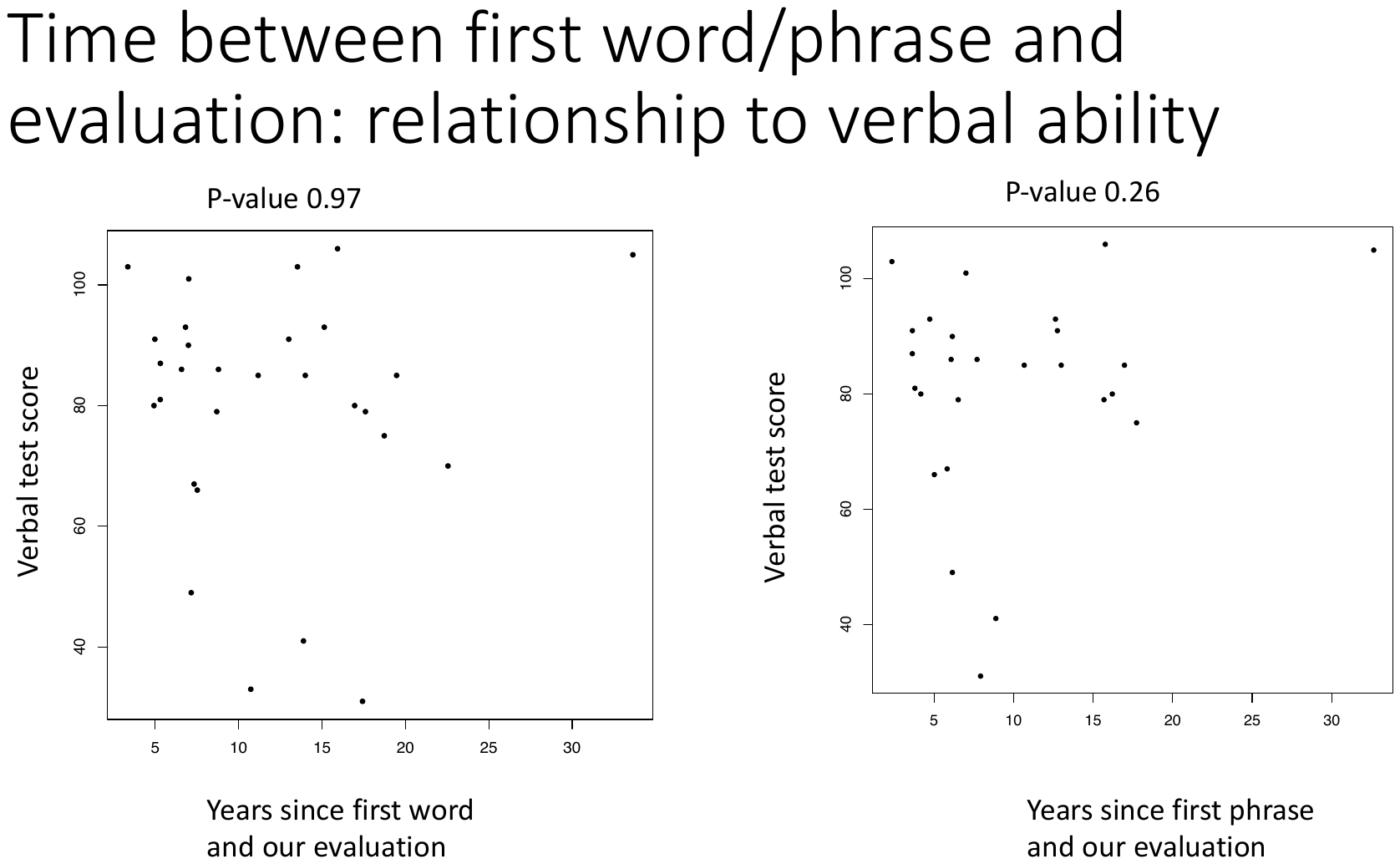

**Figure S5:** Relationship between interval (in years) from first word/first phrase and evaluation. No significant relationships were detected.
