## Supplementary material for "A distinct cognitive profile in individuals with 3q29 deletion syndrome": table 1

| **Demographic or Clinical Variable** | **mean (SD)** | **Range** |
| --- | --- | --- |
| Age in years | 14.5 (8.3) | 4.8 - 39.1 |
|  | **Percent** | **N** |
| Sex: Male | 63% | 20 |
| Ethnicity: Non-Hispanic | 97% | 31 |
| Race: White | 91% | 29 |
| Race: More than 1 Race | 9% | 3 |
| Diagnosis: ID | 34% | 11 |
| Diagnosis: ASD | 37.5% | 12 |
| Diagnosis: any anxiety disorder* | 40% | 13 |
| Diagnosis: ADHD | 63% | 20 |
| Diagnosis: Psychosis** | 19% | 4 |

**Table 1: Demographic and clinical characteristics of the 3q29 deletion study sample**

*****Anxiety disorders included generalized anxiety disorder (22%, n = 7), specific phobia (19%, n = 6), separation anxiety (12.5%, n = 4) and social anxiety disorder (6%, n = 2). 18% (n=6) had more than one anxiety disorder diagnosis.

**Psychosis evaluated in 21 subjects (psychosis not evaluated in 11 subjects; these subjects were either younger than 8 years (n = 9), or their intellectual ability or developmental level was not high enough for the evaluation to be informative (n = 2).
